# Sensitive and multiplexed RNA detection with Cas13 droplets and kinetic barcoding

**DOI:** 10.1101/2021.08.02.21261509

**Authors:** Sungmin Son, Amy Lyden, Jeffrey Shu, Stephanie I. Stephens, Parinaz Fozouni, Gavin J. Knott, Dylan C. J. Smock, Tina Y. Liu, Daniela Boehm, Camille Simoneau, G. Renuka Kumar, Jennifer A. Doudna, Melanie Ott, Daniel A. Fletcher

**Affiliations:** Department of Bioengineering, University of California, Berkeley, CA, USA; UC Berkeley-UC San Francisco Graduate Program in Bioengineering, University of California, Berkeley, Berkeley, CA, USA; Gladstone Institute of Virology, Gladstone Institutes, San Francisco, CA, USA; Medical Scientist Training Program, University of California, San Francisco, San Francisco, CA 94143, USA; Biomedical Sciences Graduate Program, University of California, San Francisco, San Francisco, CA 94143, USA; Department of Medicine, University of California, San Francisco, San Francisco, CA 94143, USA; Department of Molecular and Cell Biology, University of California, Berkeley, CA, USA; Innovative Genomics Institute, University of California, Berkeley, Berkeley, CA, USA; Monash Biomedicine Discovery Institute, Department of Biochemistry & Molecular Biology, Monash University, VIC 3800, Australia; Molecular Biophysics and Integrated Bioimaging Division, Lawrence Berkeley National Laboratory, Berkeley, CA, USA; Howard Hughes Medical Institute, University of California, Berkeley, Berkeley, CA, USA; Department of Chemistry, University of California, Berkeley, Berkeley, CA, USA; California Institute for Quantitative Biosciences (QB3), University of California, Berkeley, Berkeley, CA, USA; Division of Biological Systems and Engineering, Lawrence Berkeley National Laboratory, Berkeley, CA, USA; Chan-Zuckerberg Biohub, San Francisco, CA, USA

## Abstract

Rapid and sensitive quantification of RNA is critical for detecting infectious diseases and identifying disease biomarkers. Recent direct detection assays based on CRISPR-Cas13a^1–4^ avoid reverse transcription and DNA amplification required of gold-standard PCR assays^5^, but these assays have not yet achieved the sensitivity of PCR and are not easily multiplexed to detect multiple viruses or variants. Here we show that Cas13a acting on single target RNAs loaded into droplets exhibits stochastic nuclease activity that can be used to enable sensitive, rapid, and multiplexed virus quantification. Using SARS-CoV-2 RNA as the target and combinations of CRISPR RNA (crRNA) that recognize different parts of the viral genome, we demonstrate that reactions confined to small volumes can rapidly achieve PCR-level sensitivity. By tracking nuclease activity within individual droplets over time, we find that Cas13a exhibits rich kinetic behavior that depends on both the target RNA and crRNA. We demonstrate that these kinetic signatures can be harnessed to differentiate between different human coronavirus species as well as SARS-CoV-2 variants within a single droplet. The combination of high sensitivity, short reaction times, and multiplexing makes this droplet-based Cas13a assay with kinetic barcoding a promising strategy for direct RNA identification and quantification.

## MAIN TEXT

The PCR-based assays are currently the gold standard for RNA detection, as they can achieve high sensitivity (∼1 copy/µL) with assay times under 2 hours^6^. CRISPR-Cas13, a type VI CRISPR system, offers an alternate method for quantifying RNA by using its RNA-activated RNase activity to cleave a fluorescent reporter^7^. Though Cas13 can be combined with reverse transcription, DNA amplification, and transcription to increase sensitivity^8,9^, direct detection of RNA with Cas13 avoids the limitations of those steps and can achieve modest sensitivity by combining multiple crRNAs recognizing different regions of the target RNA. For the SARS-CoV-2 genome, LbuCas13a directly measured as low as ∼200 copies/µL in 30 minutes with 3 crRNAs in a mobile phone detector^1^ and ∼63 copies/µL in 2 hours with 8 crRNAs in a standard plate reader^2^. However, PCR-level sensitivity has not yet been achieved with direct Cas13 detection. In addition, approaches for identifying which of the multiple viral variants are present in a single sample are limited^10^.

Here we show that RNA detection with high sensitivity and multiplexed specificity can be achieved in short times by encapsulating the Cas13 reaction in droplets and monitoring enzyme kinetics using fluorescence. Like droplet digital PCR (ddPCR)^11^, our assay enables quantification of the absolute amount of target RNA based on the number of positive droplets. Unlike commercially-available ddPCR, the small droplet volume in our assay accelerates signal accumulation of the direct Cas13 reaction. When a single target RNA is encapsulated in a ∼10 pL droplet, the Cas13 signal accumulation rate is equivalent to that of a bulk reaction containing 10^5^ copies/µL target (Fig. 1A). To rapidly generate ∼10 pL droplets, we emulsified the reaction mix containing LbuCas13a for 2 min in an excess volume of an oil/surfactant/detergent mixture using an automatic multi-channel pipettor (Fig. 1B, Supplementary Fig. 1A and 1B, Methods). We imaged the resulting emulsion on a fluorescence microscope, which revealed the formation of millions of droplets, with diameters ranging from 10 to 40 µm (Fig. 1C, Supplementary Fig. 1A). Imaging the droplets allowed us to normalize the fluorescence signal by droplet size^12^ and avoid the need for slower and more complex systems to generate uniform droplet sizes.

**Figure 1:**
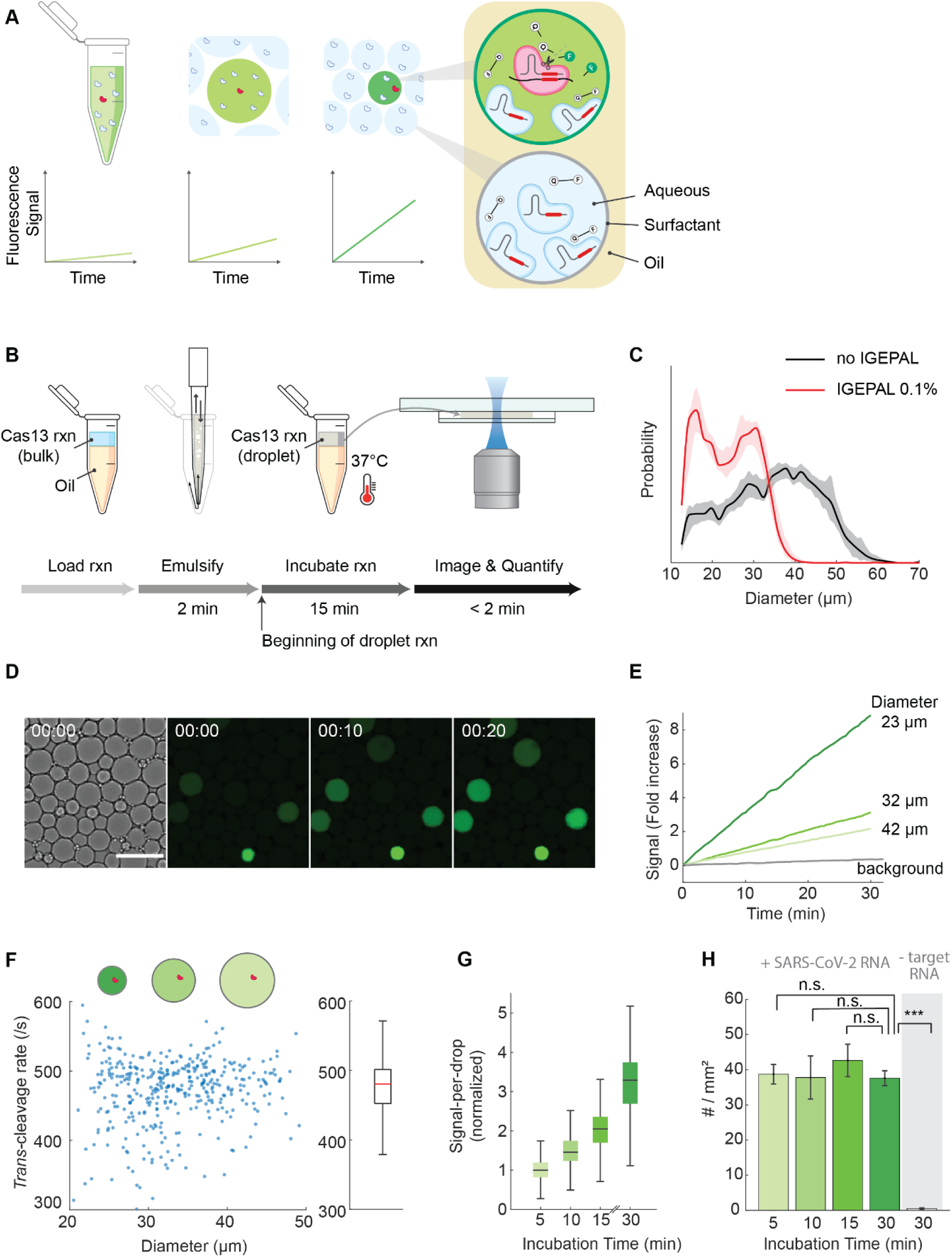
Rapid detection of the single-molecule Cas13a reaction within heterogeneous droplet. a) Schematic showing the increased signal accumulation rate for a single Cas13a confined in decreasing volumes (red: activated Cas13a, white: inactive Cas13a). Each droplet contains hundreds of thousand copies of the Cas13a-crRNA RNP (ribonucleoprotein complex) and millions of quenched RNA reporter. Only a droplet possessing one or more target RNA molecules will acquire signal. A 10pL droplet containing a single target RNA will accumulate signal at a rate equivalent to a bulk reaction containing 10^5^ copies/µL target. b) Workflow of a droplet Cas13a assay. A Cas13a reaction including target RNAs is mixed with an oil (HFE 7500 including 2% w/w Perfluoro-PEG surfactant) and emulsified by repeated pipette mixing at a constant speed for 2 minutes. The emulsified reaction is typically incubated for 15 minutes at 37°C. Subsequently, the emulsion is loaded into a custom flow cell and imaged with a fluorescent microscope. The complete assay from sample loading to imaging takes 20 minutes. c) Size distribution of droplets is reduced when 0.1% wt/vol IGEPAL is added to the Cas13a mix (the red line) compared to no IGEPAL (the black line). The shadows indicate the S.D. from 5 independent droplet preparations. d) The bright field (left) and fluorescent images of a Cas13a reaction taken with a 20X objective lens. T = 0, 10, and 20 minutes since the beginning of imaging. Scale bar = 65 µm. e) Fluorescence signal over time in three positive droplets (the green lines) and one background droplet (the grey line). Signal is the mean fluorescent intensity change within a droplet normalized by its initial value after background subtraction. An image is taken every 30 seconds and corrected for photobleaching (Methods). f) Single Cas13a turnover frequency measured from individual droplets containing crRNA 4, SARS-CoV-2 RNA, and 400nM 5U-fluorescent reporter (N = 478 droplets). The box and whisker plot in the right panel indicates the median, the lower and upper quartiles, and the minimum and the maximum values ignoring outliers. g) Signal-per-droplet with increasing incubation times for a bulk reaction containing 1 × 10^4^ copies/µL of SARS-CoV-2 RNA is obtained from positive droplets in three technical replicates. Data are represented as a box and whisker plot representing the median, the lower and upper quartiles, and the minimum and the maximum values ignoring outliers. The signal is normalized by the median signal at the 5-minute timepoint. h) Number of positive droplets with increasing incubation times are represented as mean ± SD of three technical replicates. The mean of total positive droplets is 290, 283, 320, 282, and 3.7 for 5, 10, 15, 30-minute timepoint of reaction containing target RNAs and 30-minute timepoint without any target RNA, respectively. The plotted data is represented as #/mm^2^ after dividing the total number of positive droplets by the total area imaged. P-values based on a two-tailed *Student’s* t-test are 0.61, 0.96, 0.16, and 6.86e-6, respectively. ns = not significant, ***p < 0.001.

### Droplet Cas13a assay improves the speed of direct RNA detection

We validated the Cas13 droplet assay by first forming droplets containing 10,000 copies/µL of SARS-CoV-2 RNA, LbuCas13a, crRNA targeting the SARS-CoV-2 N gene (crRNA 4) and a fluorophore-quencher pair tethered by pentauridine (5U) RNA (reporter) and then monitoring the reaction in droplets over time (Fig. 1D). At this target concentration, ∼7% of the droplets should contain the target RNA, with the vast majority of those containing only a single copy. As expected, we found that the signal accumulation rate in positive droplets was inversely proportional to droplet size (Supplementary Fig. 1C), with smaller droplets increasing faster than larger droplets (9-fold increase for a 23 µm droplet vs. 3-fold increase for a 42 µm droplet) (Fig. 1E). Our measurements show that a single LbuCas13a can cleave 471 ± 47 copies of reporter every second in the presence of 400nM reporter, or a K_cat_/K_M_ of 1.2 × 10^9^ M^-1^s^-1^, which is two orders-of-magnitude higher than that measured for LbCas12a^13^ and consistent with that measured for LbuCas13a based on a bulk assay^14^. Notably, the absolute *trans*-cleavage rate of a single LbuCas13a remains consistent regardless of droplet size (Fig. 1F, Supplementary Fig. 1D). Longer incubation times resulted in a linear increase in average signal per droplet (Fig. 1G), but all the positive reactions could be correctly identified as early as 5 minutes with a 20X/0.95NA objective (Fig. 1H) and 15 minutes with a 4X/0.20NA objective (Supplementary Fig. 1E and 1F), while the “no target” condition rarely showed positive droplets (Fig. 2).

**Figure 2:**
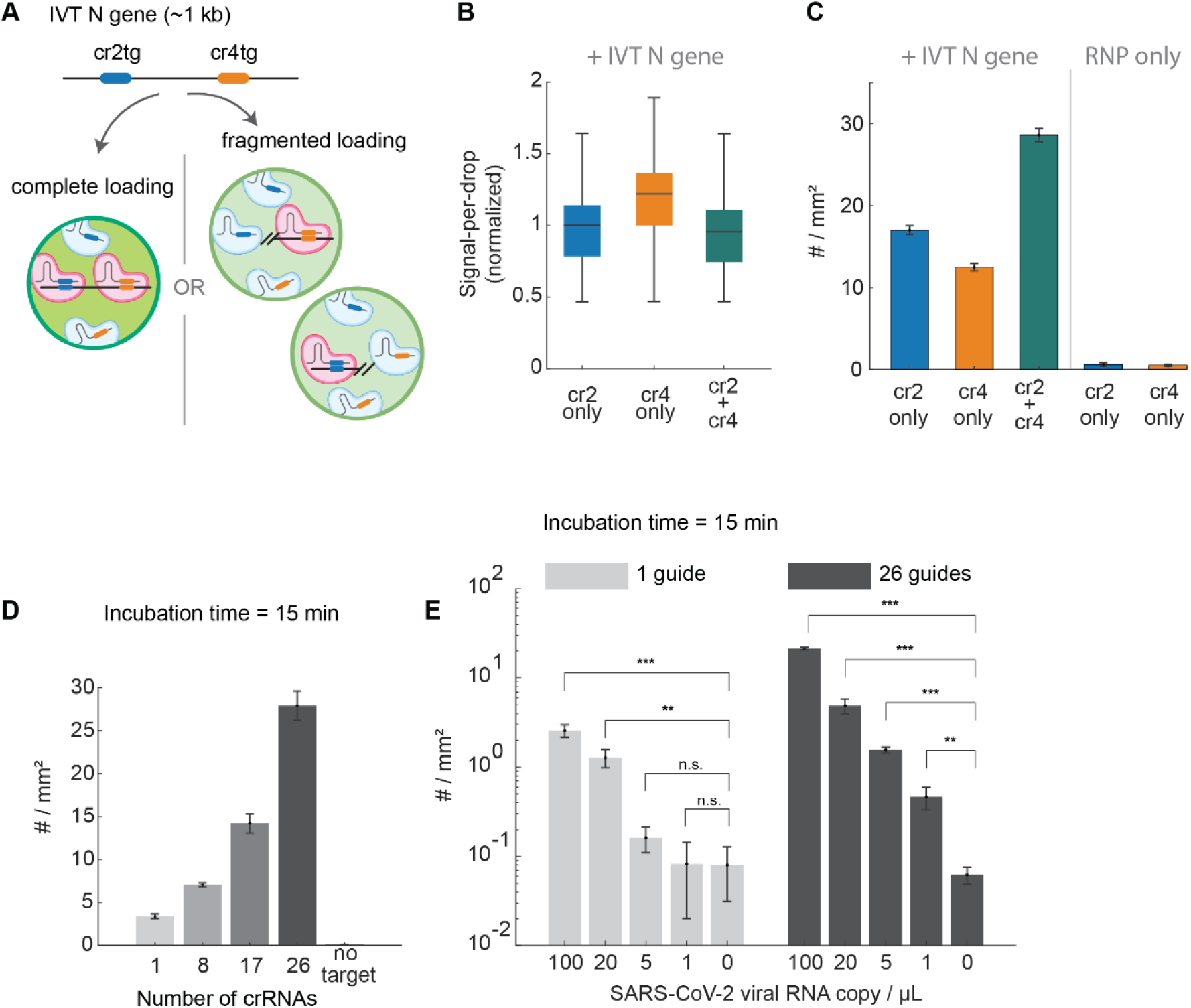
Detection sensitivity of droplet Cas13a assay using crRNA combinations. a) Schematic showing potential results of Cas13 droplet reaction combining two crRNAs. cr2tg and cr4tg depict the RNA target for the crRNA2 and crRNA4 respectively. If the whole N gene is loaded to a droplet containing both crRNAs, signal will accumulate twice as fast than the droplet containing only one target sequence (complete loading); if N gene is fragmented into two halves and loaded into two separate droplets, the number of positive droplets will be doubled while the signal of individual ones remain identical to the droplet containing one target sequence (fragmented loading). b) The result of Cas13a reaction with 2.5 × 10^4^ copies/µL of IVT N gene or without any target RNA in droplets containing crRNA 2, crRNA 4, or both crRNAs are quantified after 1 hour of reaction incubation from three technical replicates. The distribution of signal-per-droplet is represented as a box and whisker plot showing the median, the lower and upper quartiles, and the minimum and the maximum values ignoring outliers. c) As in b), but the number of positive droplets is shown. The mean of total positive droplets is 118, 90, 205, 0.67, and 0.33, for crRNA 2, crRNA 4, or both crRNAs with target RNAs and crRNA 2 and crRNA 4 without any target, respectively, and represented as mean ± SD of #/mm^2^. d) Number of positive droplets are quantified for different crRNA combinations after adding 100 copies/μL of externally quantified SARS-CoV-2 RNA (BEI resources). No target condition is measured with the 26-crRNA combination. Each reaction was incubated for 15 minutes and droplet images were taken with a 4X objective. Data are represented as mean ± SD of three replicates. e) Number of positive droplets were quantified for a dilution series of externally quantified SARS-CoV-2 RNA. Each reaction was incubated for 15 minutes. Data are represented as mean ± SD of three replicates. P-values for 100, 20, 5, and 1 target copies/µl compared to RNP only were determined based on a two-tailed *Student’s* t-test and are 5.0e-4, 2.2e-3, 0.12, 0.96, respectively, for results obtained with a single guide and 1.1e-6, 7.8e-4, 2.8e-5, 7.6e-3, respectively, for results obtained with a combination of 26 guides. ns = not significant, *p < 0.05, ** p < 0.005, ***p < 0.001.

### crRNA combinations boost the sensitivity of the droplet Cas13a assay

We next tested whether crRNA combinations, which can generate more signal per target RNA^1^, could further increase the speed of the Cas13a droplet assay. We loaded *in vitro* transcribed (IVT) target RNA corresponding to the N gene of SARS-CoV-2 to droplets containing either crRNA 2, crRNA 4, or both crRNAs, which target different regions of the N gene (Fig. 2A), and then quantified the number and signal of the positive droplets. Surprisingly, while crRNAs 2 and 4 generate similar signals when used individually (Supplementary Fig. 2A) and might be expected to double the signal when combined, we found that the signal per droplet was not significantly different in any of the positive droplets (Fig. 2B). Rather, the number of positive droplets almost doubled when droplets contain both crRNA 2 and crRNA 4 compared to just one of the crRNAs (Fig. 2C). This suggests that the N gene IVT was fragmented through *cis*-cleavage upon initiation of the reaction prior to droplet formation, causing the regions targeted by different crRNAs to be loaded into separate droplets (Fig. 1B, Fig. 2A). The resulting fragmented loading we observe is consistent with a recent study of microwell-based Cas13a reaction^3^ and suggests that multiple crRNAs activate independent Cas13a reactions whether in bulk solution or in droplets.

We then tested whether increasing crRNA combinations would further increase the speed of the Cas13a droplet assay. We identified 26 crRNAs targeting various regions of the SARS-CoV-2 genome that individually produce a strong signal in Cas13 detection reactions (Supplementary Table 1). Adding more crRNAs while keeping the total RNP concentration constant led to an increasing number of positive droplets (Fig. 2D). This was possible because the activity of Cas13a remains constant even when only a small fraction (1/50 or less) of total RNPs in a droplet contains crRNAs matching the target (Supplementary Fig. 2B). Overall, this suggests that a large number of crRNAs can be combined to maximize the number of independent Cas13a reactions from the same target RNA. Given that the sensitivity of droplet-based assays is fundamentally limited by the false-positive rate observed in the absence of target RNA, the generation of multiple positive droplets per target RNA has the potential to increase the signal-to-background, and thus, sensitivity of the assay.

To test the sensitivity of our Cas13a droplet assay with crRNA combinations, we carried out serial dilutions of precisely quantified SARS-CoV-2 genomic RNA obtained from the Biodefense and Emerging Infections Research Resources Repository (BEI Resources). We quantified the number of positive droplets in each dilution using either a single crRNA or all 26 crRNAs, using 36 images per condition (∼160,000 droplets) after 15 minutes of reaction incubation (Fig. 2E). For the single crRNA (crRNA 4), the number of positive droplets was significantly higher than the no-target control for the samples containing 20 target copies/µL or more. For the combination of 26 crRNAs, the limit of direct detection was 1 copy/µL target, comparable to the sensitivity of PCR. This limit of detection was not improved if we incubated the reaction for 30 minutes instead of 15 minutes (Supplementary Fig. 2C).

### crRNA/target RNA combinations govern Cas13a reaction kinetics

The fast Cas13a kinetics we observe in droplets critically depends on the choice of the crRNA and its target. For example, two other crRNAs targeting the SARS-CoV-2 N gene—crRNA 11 and crRNA 12—exhibit significantly slower rates in bulk reactions than crRNA 2 or 4 (Fig. 3A, Supplementary Fig. 2A). For this reason, the selection of “good” crRNAs that support efficient Cas13 activity is critical for bulk Cas13-based molecular diagnostics^15^, though how different crRNAs affect the activity of Cas13 is not well understood^16^. We therefore tested crRNAs 11 and 12 in our droplet assay and compared it to the activity of crRNA 4. As expected, the number of positive droplets was reduced for crRNAs 11 and 12, but the difference to crRNA 4 was less than observed in bulk reactions (Fig. 3B). On the other hand, the signal in each positive droplet was significantly reduced for crRNA 11 and 12 compared to crRNA 4 (Fig. 3C). To better understand how different crRNAs affect Cas13a activity, we examined the individual reaction trajectories within positive droplets, which report the change in fluorescence between each 30 second measurement time point (Fig. 3D-F). For all three crRNAs, most positive droplets exhibited slopes significantly higher than those recorded in RNP-only droplets lacking the target RNA (Fig. 3G). However, the three crRNAs induced distinct kinetic behaviors in droplets, with the slope, shape, and x-intercept of the individual trajectories varying widely depending on the individual crRNA. This ultimately resulted in different endpoints for each crRNA. In some cases, we found a strikingly stochastic behavior in which trajectories exhibited periods of no signal increase followed by periods of rapid signal increase (“rugged” vs smooth trajectory) (Fig. 3H-I). Since each droplet contains, on average, a single target, these results indicate that specific crRNA/target combinations modulate Cas13a enzymatic activity at a single-molecule level.

**Figure 3:**
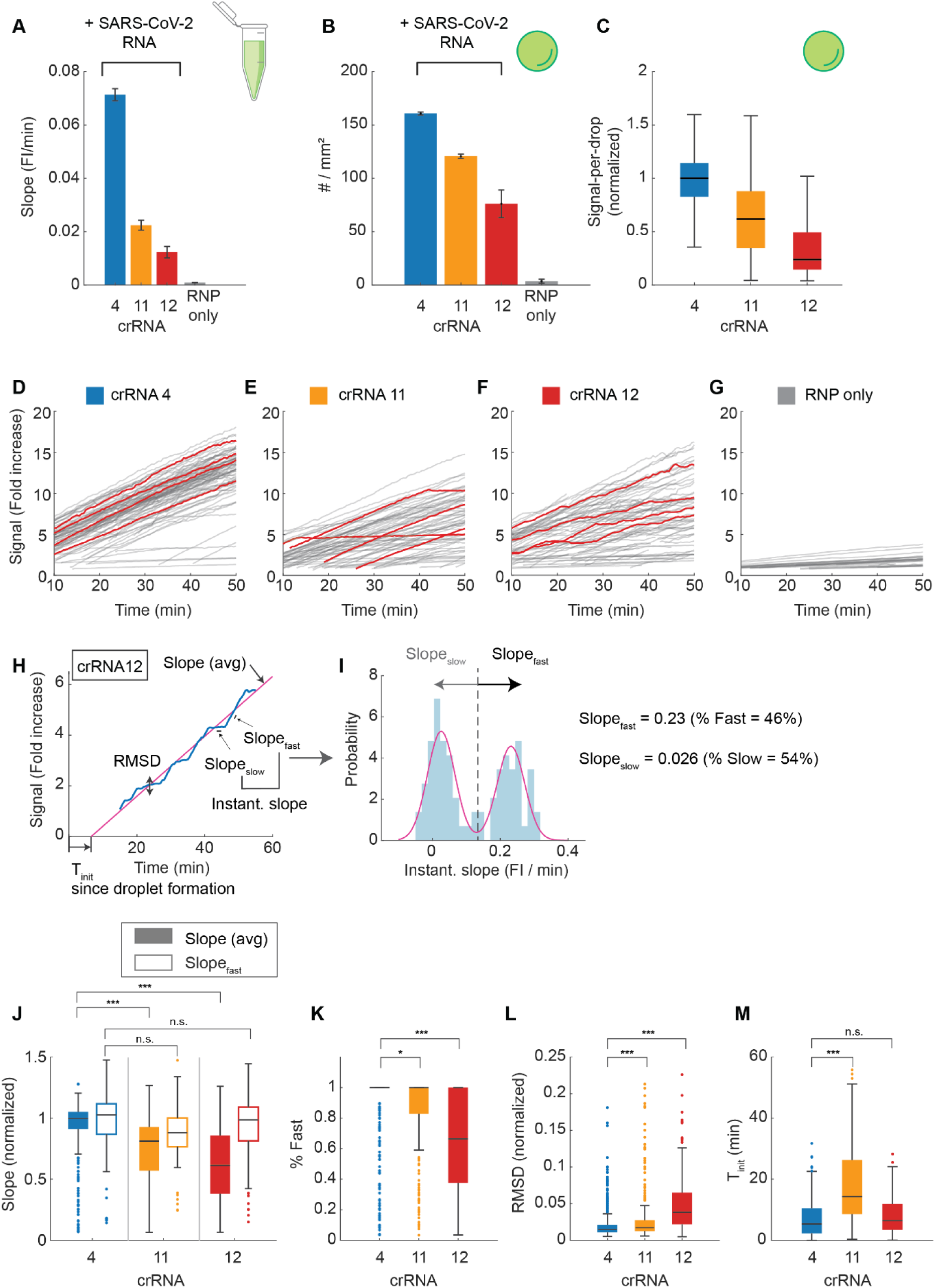
The crRNA-dependent heterogenous Cas13a activities. a) The slope of a bulk Cas13a reaction with 3.5 × 10^4^ copies/µL of SARS-CoV-2 RNA or without any target (RNP only) for crRNAs targeting different regions of SARS-CoV-2 N gene. RNP only condition is measured with all three crRNAs. The slope is determined by performing simple linear regression of data from each replicate (N = 3) individually. Data are represented as mean ± SD. b) The result of droplet Cas13a reaction with 3.5 × 10^4^ copies/µL of SARS-CoV-2 RNA or without any target (RNP only) after 30 minutes of incubation. A total of 1287, 966, 605, and 21 positive droplets were quantified from reactions using crRNA 4, crRNA 11, crRNA 12 and RNP only, respectively, and shown as #/mm^2^. Data are represented as mean ± SD of three replicates except for crRNA 4, where two replicate experiments were performed. The RNP only condition is measured with all three crRNAs combined. c) As in b), but the signal-per-droplet for different crRNAs are represented as a box and whisker plot showing the median, the lower and upper quartiles, and the minimum and the maximum values ignoring outliers. Signal is normalized by the median of crRNA 4. d) Time trajectories of single Cas13a reactions using crRNA 4. 100 individual trajectories are measured in droplets ranging from 30 to 36 µm in diameter (grey lines) and representative trajectories are shown as red lines. Signal is measured every 30 seconds and represented as fold increase from the initial value. Two replicate experiments were combined for each crRNA. e) As in d), but with crRNA 11. f) As in d), but with crRNA 12. g) Time trajectories of the rare positive droplets measured from a Cas13a reaction containing an 8-crRNA combination (Supplementary Table 1) and no target RNA. 31 individual trajectories from two replicate runs were measured in droplets ranging from 30 to 36 µm in diameter. h) Analysis strategy of individual Cas13a signal trajectories. The blue curve is a crRNA 12 example trajectory. The average slope (Slope (avg)), T_init_, and the Root-mean-square-deviation (RMSD) are determined by performing simple linear regression to the raw signal. Slope_fast_ and Slope_slow_ correspond to the fast and slow periods determined in i). i) Instantaneous slopes are calculated by taking the time-derivative of the raw signal (the blue histogram) and fitting its probability distribution with either single- or binary-Gaussian (the red line). For a data that favors binary distribution, we determined Slope_fast_, Slope_slow_, % Fast, and % Slow by taking the mean and the proportion of each Gaussian. j) Distribution of slope is represented as a box and whisker plot showing the median, the lower and upper quartiles, and the minimum and the maximum values, with outliers overlaid as individual data points. Individual 30-minutes-long trajectories from droplets of arbitrary size are used after their signal is normalized for droplet size. 395, 267, and 311 trajectories are used for crRNA 4, crRNA 11, and crRNA 12, respectively. P-values determined from a two-tailed *Student’s* t-test are 2.1e-20, 7.1e-45, 0.34, 0.39. ns = not significant, *p < 0.05, ***p < 0.001. k) As in j), but for % fast. P-values determined from a two-tailed *Student’s* t-test are 0.021 and 4.3e-34. l) As in j), but for RMSD. P-values determined from a two-tailed *Student’s* t-test are 1.5e-4 and 4.4e-30. m) As in j), but for T_init_. P-values determined from a two-tailed *Student’s* t-test are 1.6e-23 and 7.7e-2.

To quantify these kinetic differences in droplets, we characterized individual trajectories by their average slope, root-mean-square-deviation (RMSD), and time from target addition to initiation of enzyme activity (T_init_) (Fig. 3H). By calculating the instantaneous slopes at each point in the trajectory and fitting their distribution to a Gaussian, we found that the trajectories exhibited two different slopes, one “fast” when fluorescence was increasing and one “slow” when fluorescence was not increasing (Fig. 3I). Interestingly, even though the average slopes differed significantly for the different crRNAs, the instantaneous “fast” slopes were constant across all three crRNAs (Fig. 3J). Consistent with this, crRNA 12, which exhibited the lowest average slope of the three crRNAs, showed extended “slow” periods (Fig. 3K) and increased level of signal fluctuation (Fig. 3L). We found that T_init_ varied significantly among the three crRNAs we tested, with crRNA 11 exhibiting the slowest T_init_ (Fig. 3M), resulting in reduced signal at the reaction endpoint (Fig. 3C). To test if the stochastic behavior of individual Cas13a reactions is simply caused by the unbinding of crRNAs from the Cas13a protein, we changed the RNP concentration to below or above the K_d_ of the crRNA-Cas13a complex^7,17^. The stochastic behavior was unaltered (Supplementary Fig. 3A and 3B) and crRNA 4 and crRNA 12 retained their characteristic kinetic features even when droplets contained only single copies of Cas13a and crRNA (Supplementary Fig. 3C). In contrast, when we changed the target from genomic SARS-CoV-2 RNA to a 20-nucleotide fragment complementary to crRNA 12’s spacer sequence, the stochastic behavior of the reaction was no longer observed and T_init_ was significantly shortened (Supplementary Fig. 4). This suggests that RNA regions outside of the targeted spacer sequence affect Cas13a activity.

### Heterogenous Cas13a kinetics offer a method for multiplexed detection

We hypothesized that the distinct kinetic signatures of the different crRNAs could be used to identify specific crRNA-target pairs in one droplet, thus providing a method for multiplexed detection of different RNA viruses (Fig. 4A) or variants of the same virus (Fig. 4B). To test this idea, which we termed ‘kinetic barcoding’, we combined a crRNA targeting a common cold virus HCoV-NL-63 (HCoV crRNA 6) and a second crRNA targeting SARS-CoV-2 (crRNA 12); both crRNAs were chosen because they individually exhibit different kinetic signatures on their respective targets (Fig. 4C). We collected 30-minute trajectories from hundreds of droplets containing either HCoV-NL-63 or SARS-CoV-2 viral RNA along with Cas13a and both crRNAs together. The two groups of trajectories associated with the two different crRNAs were clearly distinguishable based on their average slope and RMSD (Fig. 4D). To determine how clearly those two groups can be distinguished, we randomly sampled a subset of trajectories and compared their difference by performing Student’s t-test on their binary classification result (Fig. 4E) (see Methods). As expected, we found that increasing the number of trajectories and extending the measurement time improved classification (Supplementary Fig. 5A), though measurement times longer than 10 minutes did not further improve the classification. This analysis showed that kinetic barcoding can distinguish between HCoV-NL-63 and SARS-CoV-2 targets within 10 minutes provided that 20 or more trajectories are measured; no difference in performance was observed when images were acquired every 3 minutes for a 30-minute period or every 30 seconds for a 10-minute period (Supplementary Fig. 5B).

**Figure 4:**
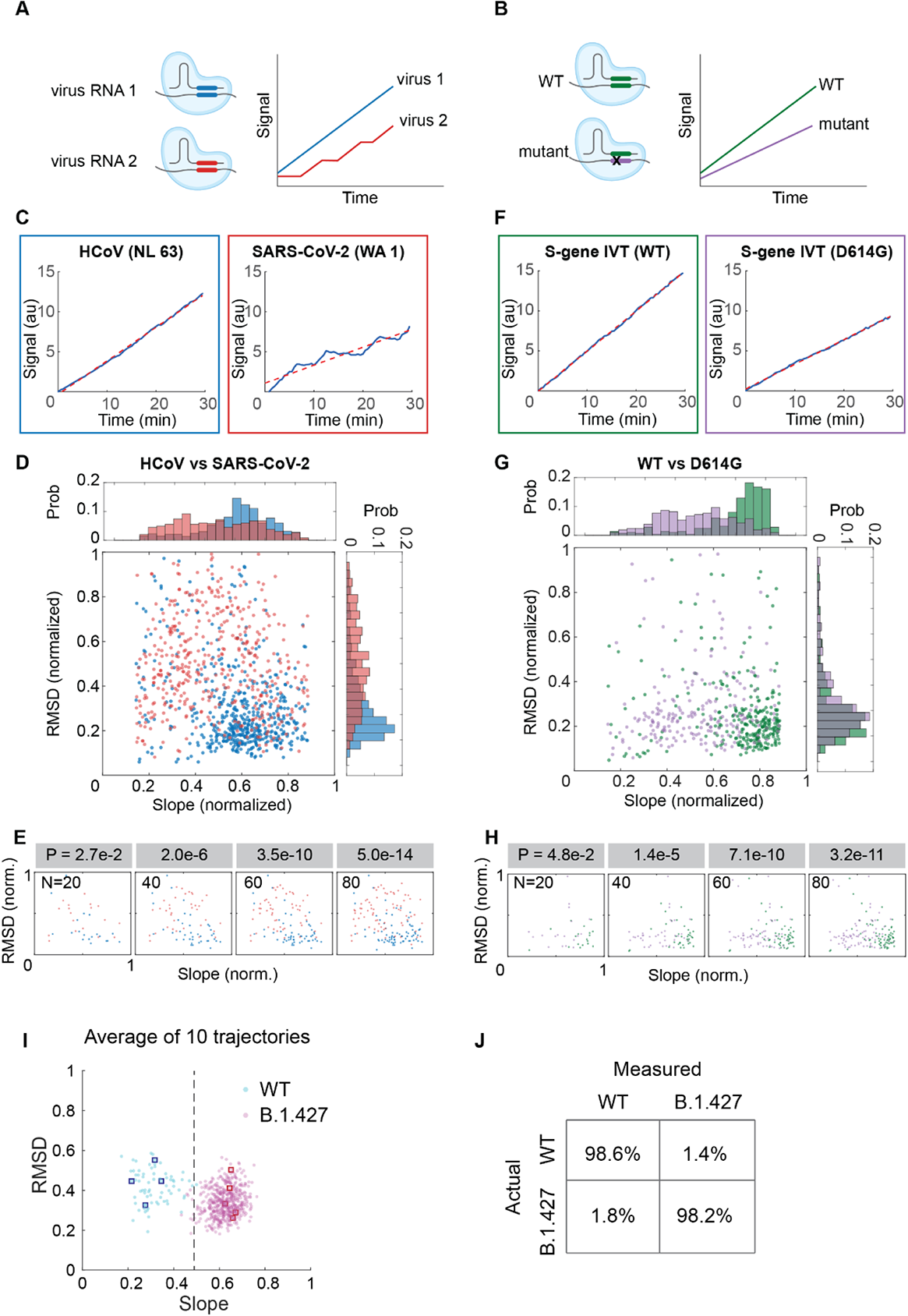
The kinetic-barcoding approach for multiplexed detection of viruses or virus variants. a) Schematic showing the principle of kinetic-barcoding approach for simultaneous detection of two different viruses. It relies on the unique Cas13a kinetic signatures for a specific combination of crRNA and its target. b) As in a), but detection of two different variants. c) Representative single Cas13a reaction trajectories with HCoV-NL-63 RNA (targeted by crRNA 7) or SARS-CoV-2 RNA (targeted by crRNA 12). Signal is the total fluorescence change in a droplet normalized by its initial value, which remains invariant regardless of droplet size. The dotted red line shows a linear fit. d) Distribution of slope and RMSD of individual Cas13a trajectories between HCoV-NL-63 and SARS-CoV-2 RNA. The slope and RMSD is determined from individual 30 minutes-long trajectories (N = 488) measured with a single crRNA-target pair. The RMSD is first normalized by the mean signal of the same trajectory and then normalized to 0 to the highest value of the dataset. The slope is normalized to 0 to the highest value in the dataset (blue = HCoV-NL-63; red = SARS-CoV-2). e) Identification of HCoV-NL-63 or SARS-CoV-2 based on the kinetic parameters of individual Cas13a reactions. Varying numbers of 30-minute-long Cas13a trajectories are randomly selected from each condition and the difference between two groups are quantified as p-values based on a two-tailed *Student’s* t-test. f) Representative single Cas13a reaction trajectories with the WT SARS-CoV-2 S gene or the one containing the D614G mutation. The dotted red line shows a linear fit. g) Distribution of slope and RMSD of individual Cas13a trajectories between the WT and D614G mutant (N = 208). h) As in e), measured for WT vs D614G. i) Detection of SARS-CoV-2 variant from clinical samples based on kinetic-barcoding. The average of the slope or RMSD distribution is obtained by randomly selecting 10 positive trajectories from each sample (the blue dots are WT representing 69 independent trials and the magenta dots are B.1.427 representing 494 independent trials). The blue and magenta squares indicate an example values for each sample. The black dotted line indicates the slope threshold (Slope = 0.49) separating the WT from B.1.427. j) Detection specificity of kinetic barcoding. The confusion matrix is determined from i).

Similar experiments were performed to differentiate the original WA1 SARS-CoV-2 strain from common variants. First, we used a crRNA (crRNA 6) spanning the wild-type D614 sequence in the SARS-CoV-2 Spike (S) gene and compared the signal trajectories generated from IVT wild-type S gene with the trajectories from the S gene encoding a D614G mutation now shared by most SARS-CoV-2 variants since June 2020^18^ (Fig. 4F). Both crRNAs yielded predominantly “smooth” trajectories (as indicated by the low RMSD), but the average slopes obtained with the mutant target were significantly lower than those obtained with the matching wild-type target (Fig. 4G). Based on this difference in slopes, we distinguished the D614G mutant from the wild-type target within 5 minutes using a minimum of 30 trajectories (Supplementary Fig. 5C).

Second, we focused on a specific California variant of concern (B.1.427/B.1.429; Epsilon), which among others harbors a unique S13I mutation in the S protein; this variant exhibits ∼20% increased transmissibility and a 4- to 6.7-fold and 2-fold reduced neutralization by convalescent and post-vaccination sera, respectively^19^. We used a crRNA matching the mutant sequence (crRNA 45) and measured RNA extracted from cultured wild-type or mutant viruses as well as nasal/oral swab samples from people known to be infected with wild-type virus or the variant based on sequencing (Supplementary Table 2). The Ct values from quantitative PCR measurements in patient-derived samples were between 15 and 20 and provided 15 to 350 positive trajectories among the droplets measured. Although individual trajectories from each sample exhibited heterogeneous slopes and RMSDs, the average slopes measured in the wild-type samples were significantly lower than those measured from B.1.427 (Fig. 4I and Supplementary Fig. 5D). To test if B.1.427 could be correctly identified when only 10 individual trajectories were collected, we randomly sub-sampled 10 trajectories from each sample (Fig. 4I). We found that, regardless of the choice of trajectories, the average slope distribution was clearly distinguishable between wild-type virus and the B.1.427 variant, providing a detection accuracy of >98% for both cases (Fig. 4J).

### Amplification-free Cas13 reactions in droplets enable sensitive, multiplexed RNA detection

In summary, we demonstrate that a droplet-based Cas13a direct detection assay can achieve PCR-level sensitivity and distinguish RNA targets based on reaction kinetics. As one crRNA can be diluted at least 50 times or more without compromising its performance (Supplementary Fig. 2B), we speculate that combining multiple different crRNAs can be used to further enhance detection sensitivity and bring the limit of detection below 1 copy/µL. At this sensitivity, a droplet-based Cas13a direct detection assay would be used to measure very low viral loads such as at the beginning or end of an infection, in environmental samples, or with latent viruses such as HIV, without the need for additional reverse transcription or amplification steps.

We found that LbuCas13a is an efficient, diffusion-limited enzyme, whose kinetics are controlled by the specific combination of the crRNA and the target^20,21^. Interestingly, the distribution of Cas13a RNP trajectories is largely homogenous for “good” crRNAs supporting high activities (Fig. 1F), implying that the active conformation of Cas13a RNP is stable over time. However, certain “bad” crRNAs were found to induce “rugged” trajectories and to switch off Cas13a activity for more than a minute. The observation that this kinetic feature is abolished when a short RNA fragment is present instead of the viral RNA (Supplementary Fig. 4) suggests a role of local or global RNA structure or its chemical modifications in the kinetics of an individual Cas13a RNP – features that are lost during target amplification – rather than an effect of enzyme conformational switching^22^. On the other hand, we show that sequence mismatches between the crRNA and its target can reduce the slope of a reaction without introducing “rugged” stochastic activity switching (Fig. 4G and H), pointing to multifactorial mechanisms governing Cas13a kinetics. Taking advantage of these characteristic kinetic signatures, we are able to conclusively determine which virus or variant was present in a given droplet, demonstrating the diagnostic utility of kinetic barcoding and establishing kinetic variety as an additional feature to be considered during crRNA selection.

Digital assays enhance the sensitivity and quantitative performance in ddPCR^23,24^, protein detection^25^, and recently CRISPR-Cas-based nucleic acid detection approaches^3,4,26,27^. However, commercially-available droplet technologies tailored for ddPCR assays were not suitable for this assay, as amplification-free Cas13a assays require smaller droplets (∼10pL) than those typically used in ddPCR (∼900pL^28^) to achieve the desired signal accumulation. On the other hand, microwell-based assays often encapsulate reactions in femtoliter-scale volumes^3^), reducing the sample volume and limiting the detection sensitivity. We show that for a compartmentalized assay with average Cas13a kinetics, a ∼10pL reaction volume provides optimal speed and sensitivity. Our current workflow based on bulk droplet formation followed by direct imaging was chosen to maximize throughput at the same time as accounting for droplet heterogeneity, an approach that could be scaled up to process samples in parallel. As a laboratory assay, Cas13a reactions in droplets have potential advantages over PCR for RNA quantification by avoiding errors associated with reverse transcription and amplification, though large-scale testing of diverse samples would be needed to properly compare the two approaches. In conclusion, droplet-based Cas13a direct detection assays with kinetic barcoding represents a method for using fundamental properties of CRISPR-Cas13 reactions at a single-molecule level to enable rapid and sensitive RNA quantification that could be extended to multiple RNA viruses and RNA biomarkers in the future.

## Data Availability

The data that support the findings of this study are available from the corresponding author upon reasonable request.

## METHODS

### Protein purification

Protein purification was performed as previously described^1^. Briefly, the LbuCas13a expression vector contains the codon-optimized Cas13a genomic sequence, N-terminal His6-MBP-TEV cleavage site sequence, and a T7 promoter binding sequence (Addgene Plasmid #83482). The protein is expressed in Rosetta 2 (DE3) pLysS E. coli cells in Terrific broth at 16°C overnight. Soluble His6-MBP-TEV-Cas13a was isolated over metal ion affinity chromatography and the His6-MBP tag was cleaved with TEV protease at 4°C overnight. Cleaved Cas13a was loaded onto a HiTrap SP column (GE Healthcare) and eluted over a linear KCl (0.25-1.0M) gradient. Cas13a-containing fractions were further purified via size-exclusion chromatography on a S200 column (GE Healthcare) in gel filtration buffer (20 mM HEPES-K pH 7.0, 200 mM KCl, 10% glycerol, 1 mM TCEP) and were subsequently flash frozen for storage at -80°C.

### Preparation of SARS-CoV-2 RNA segments

*In vitro* RNA transcription is performed as previously described^1^. SARS-CoV-2 N gene, S gene (WT), and S gene with D614G mutation were transcribed off a single-stranded DNA oligonucleotide template (IDT) using HiScribe T7 Quick High Yield RNA Synthesis Kit (NEB) following manufacturer’s recommendations. Template DNA was removed by addition of DNase I (NEB), and IVT RNA was subsequently purified using RNA STAT-60 (AMSBIO) and the Direct-Zol RNA MiniPrep Kit (Zymo Research). RNA concentration was quantified by Nanodrop and copy number was calculated using transcript length and concentration.

### Preparation of full viral genomic RNA

Full viral genomic RNAs are purified as previously described^1^. Isolate USA-WA1/2020 of SARS-CoV-2 (NR-52281 BEI Resources) and CA427 (B.1.427, received from CA DPH) were propagated in Vero CCL-81 cells. Isolate Amsterdam I of HCoV-NL-63 (NR-470, BEI Resources) was propagated in Huh7.5.1-ACE2 cells (all virus cultures were performed in a Biosafety Level 3 laboratory). RNA was extracted from the viral supernatant via RNA STAT-60 (AMSBIO) and the Direct-Zol RNA MiniPrep Kit (Zymo Research).

### SARS-CoV-2 patient samples

Patient samples were obtained from a UC Berkeley campus testing program, as previously described^2^. Briefly, human mid-turbinate nasal/oropharyngeal swabs were collected, processed, and tested by running qRT-PCR on extracted RNA using primers for SARS-CoV-2. Ct values were obtained using primers for the N gene, S gene, and Orf1ab of the SARS-CoV-2. When the Ct value for two out of the three genes was 37 or below, the sample was considered positive. Ct values for the samples used in this study can be found in Supplementary Table 2. Sequence data for each sample was used to identify SARS-CoV-2 variants. The samples used in this study were non-identifiable biospecimens, for which there was no link to identifiable subject information. Only secondary data analysis was carried out on the samples, and the research data will not be held for inspection by nor submitted to the FDA.

### crRNA design

All crRNAs targeting SARS-CoV-2, SARS-CoV-2 variants (D614G, S13I), or HCoV-NL-63 were ordered as desalted oligonucleotides from Synthego at a scale of 5 nmole (Supplementary Table 1). Select crRNAs have also been previously described^1,3^.

### Bulk Cas13a nuclease assays

LbuCas13a-crRNA RNP complexes were first preassembled at 133nM equimolar concentration for 15 minutes at room temperature and then diluted to 25 nM LbuCas13a in cleavage buffer (20 mM HEPES-Na pH 6.8, 50 mM KCl, 5 mM MgCl2, and 5% glycerol) in the presence of 400 nM of reporter RNA (5’-Alexa488rUrUrUrUrU-IowaBlack FQ-3’), 1 U/μL Murine RNase Inhibitor (NEB, Cat# M0314), 0.1 vol% IGEPAL 630 (Fisher, Cat# ICN19859650), and varying amounts of target RNA. For reactions using more than one crRNA, multiple crRNAs were combined at an equal concentration and subsequently the total crRNA mix was assembled with Cas13a at 133nM equimolar concentration. 25nM of RNP complex is used unless specified otherwise. The reaction mix is measured either in bulk or in droplet following emulsification (see droplet formation); For the bulk Cas13a assay, the reaction mix was loaded into a 0.2mL 8-tube strip (Fisher Cat# 14-222-251) and incubated in a compact fluorescence detector (Axxin, T16-ISO) for 1 hour at 37°C with fluorescence measurements taken every ∼30 seconds (FAM channel, gain 20). Fluorescence values were normalized by the values obtained from reactions containing only reporter and buffer.

### Droplet formation

To emulsify the Cas13a reaction mix, 20 µL of the aqueous mix was combined with 100 µL of HFE-7500 oil supplemented with 2% (w/w) PEG-PFPE amphiphilic block copolymer surfactant (008-Fluoro-surfactant, RAN Biotechnologies) in a 0.2 mL 8 tube-strip. The oil/aqueous mix was emulsified by repeated, fully-automated pipetting using an electronic 8-channel pipette (Integra-biosciences, Part # 4623) fitted with a 200 µL pipet tip (VWR Cat # 37001-532). A sample volume of 110 µL was mixed for 150 repetitions at the maximum speed (speed 10) to generate droplets of a narrow size range. The emulsion was either directly loaded into a flow cell for the time course imaging or incubated in a heating block at 37°C before being transferred and imaged. In cases where imaging could not be performed immediately after the reaction, the emulsion was quenched on ice until imaging. In both cases, the emulsion was quickly separated by spinning in a speed-controlled mini-centrifuge (∼50 rpm) for 10 seconds, the oil was completely removed from the bottom of the tube, and the emulsion was transferred into a custom flow cell after several cycles of gentle manual mixing.

### Flow cell for droplet imaging

The sample flow cell was prepared by sandwiching double-sided tape (∼20µm thick, 3M Cat# 9457) between an acrylic slide (75mm x 25mm x 2mm, laser cut from a 2mm-thick acrylic plate) and a siliconized coverslip (22mm x 22mm x 0.22mm, Hampton research Cat# 500829). Both surfaces are hydrophobic, promoting thin layers of oil between droplets and the surface. Siliconized coverslips were rinsed with isopropanol to remove any auto-fluorescent debris (20 minutes sonication) and spin dried prior to assembly. 15 µL of sample emulsion is loaded into the flow cell by capillary action, after which the inlet and outlet are sealed with Valap (1:1:1 vasoline:lanoline:paraffin).

### Microscopy and data acquisition

Droplet imaging is carried out on an inverted Nikon Eclipse Ti microscope (Nikon Instruments) equipped with a Yokogawa CSU-X spinning disk. A 488-nm solid state laser (ILE-400 multimode fiber with BCU, Andor Technologies) is used to excite the RNA probe and the fluorescence light was spectrally filtered with an emission filter 535/40nm (Chroma Technology) and imaged on an sCMOS camera (Zyla 4.2, Andor Technologies). A 20x water-immersion objective (CFI Apo LWD Lambda S, NA 0.95) is used with the Perfect Focus System to monitor droplets in the course of reaction or to accurately quantify fluorescence signal at reaction endpoint. Images are acquired through Micro-Manager with 500ms exposure time and 2×2 camera binning. Typically, 16 field-of-views (FOV) are acquired every 30 seconds for the time course imaging and 36 FOVs are acquired for the endpoint imaging. A 4x objective (CFI Plan Apo Lambda, NA 0.20) is used for the high-throughput droplet imaging at reaction endpoint. 36 FOVs are acquired with 3s exposure time without camera binning.

### Image analysis – droplet detection

We used MATLAB (Mathworks R2020b) to detect positive droplets and quantify fluorescence signals from microscopy images. First, the grayscale images were converted to binary images based on a locally adaptive threshold. The threshold is defined generously to select all the positive droplets and potentially some negative droplets or debris at this stage. Second, connected droplets were separated by watershed transform. Third, individual droplets are identified by looking for circular continuous regions and droplet parameters such as radius, circularity, and fluorescence signal are quantified. Fluorescence signal is quantified in two different ways: the mean fluorescence signal of a droplet reflecting the density of cleaved reporter; the total fluorescence signal reflecting the total amount of cleaved reporter within a droplet. Lastly, positive droplets are chosen based on their circularity and total fluorescence signal by applying a threshold that was consistently used throughout the experiments.

### Image analysis – droplet tracking in time course images

To quantify signal accumulation in the same droplet over time, we associated droplets over time based on their motion estimated by a Kalman filter in MATLAB. The filter is used to predict the track’s location in each frame and determine the likelihood of each detection being assigned to each track. Only the droplets showing continuous trajectories in time and magnitude are selected for the downstream analysis.

### Comparison of single Cas13a reaction with enzyme kinetics

We analyzed the Cas13a reaction with a single crRNA (Fig. 1F) using the Michaelis-Menten enzyme kinetics model with the quasi-steady-state approximation:

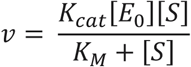

where *v* is the reaction rate, [*E*_0_] is ternary Cas13a, [*S*] is the RNA reporter, *K*_*cat*_ and *K*_*M*_ are the catalytic rate constant and the Michaelis constant. We assumed low substrate concentration ([*S*] << *K*_*M*_) since the RNA reporter [*S*] is 400nM and *K*_*M*_ is estimated to be larger than 1µM^4^ and simplified the equation to:

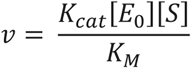

*v* /[*E*_0_] is turnover frequency, or the reciprocal of the mean waiting time <1/t> in the single-molecule Michaelis-Menten framework^5^, and it can be obtained from Supplementary Fig. 1C and D after converting the fluorescence signal to molar concentration of cleaved reporter based on a calibration.

### Data analysis – Cas13a time trajectories

We processed the raw signal in a series of steps prior to analysis: First, we corrected for the global signal fluctuation, which arise from a slight drift in z-focus even with the Perfect Focus System; we characterized the global signal from the background droplets as the mean of their pixel values and divided it from the positive droplet signal in each image frame. Second, we corrected for the photobleaching; we first characterized the signal decay rate from >200 Cas13a curves exhibiting negative slopes and positive initial signals. We modeled photobleaching as a linear function of initial signal based on the observed linear relationship between the decay rate versus initial signal (R^2^ = 0.87). Using the model, we corrected for photobleaching in each trajectory point-by-point. Third, we filtered the trajectories with a weak Savitzky-Golay filter (order 5, frame length 9) to remove the high frequency measurement noise while preserving overall structure of the curve. Lastly, we calculated instantaneous slopes by dividing signal change between frames by the frame interval and removed single outliers exhibiting high positive or negative slopes. To characterize key parameters of Cas13a kinetics, we analyzed individual trajectories in two different domains: First, we determined the slope, T_init_, and RMSD from signal time trajectories by linear regression. Since T_init_ indicates time since the droplet reaction was started, we added a constant time (12.5 minutes) that took from Cas13 droplet formation until the beginning of time course imaging. Second, we determined the slope_fast_, slope_slow_, and a fraction spent in each period by fitting a gaussian pdf to the instantaneous slope distribution. We compared the model qualities between the single versus binary gaussian pdfs using Akaike’s Information Criterion (AIC) to determine whether a trajectory exhibits two different periods of slope of not.

### Data analysis – kinetic barcoding

We used the slope and RMSD of individual signal trajectories to compare Cas13a reactions between different target-crRNAs. We first performed binary classification of trajectories based on the Supported Vector Machine (SVM) in MATLAB. For this, we collected 200 to 400 signal trajectories in each condition, in two or more independent experiments per condition to prevent bias. We converted the trajectories into a 2D array consisting of the slope and RMSD, and divided the array into a training and a validation set. We then trained an algorithm using the training set with the known answers (i.e. target-crRNA condition) and classified the validation set. The accuracy of identifying individual trajectories were 75% for HCoV-NL-63 vs SARS-CoV-2 and 73% for WT vs D614G. To access significance between two groups of trajectories, we employed a two-tailed Student’s t-test to the predicted class and reported p-values.

## ACKNOWLEDGEMENTS

We thank all members of the Fletcher, Ott, and Doudna laboratories for helpful discussions and feedback on this project. We also thank Stacia Wyman and Bryan Bach for providing samples from the Innovative Genomics Institute, and Chaz Langelier for providing samples from the Chan-Zuckerberg Biohub. Purified LbuCas13a was a kind gift from Shanghai ChemPartner, and we thank Synthego for support with synthetic crRNAs. We gratefully acknowledge support from NIH/NIAID grant 5R61AI140465-03 to J.A.D., D.A.F., and M.O. and a generous gift from an anonymous donor in support of the ANCeR diagnostics consortium, as well as generous individual donors to the Gladstone Institutes. This work was supported in part by the Health Tech CoLab at the Blum Center for Developing Economies at UC Berkeley. G.J.K. is supported by an NHMRC Investigator Grant (EL1, APP1175568). J.A.D. is an HHMI Investigator. D.A.F. is a Chan-Zuckerberg Biohub Investigator. The following reagents were deposited by the Centers for Disease Control and Prevention and obtained through BEI Resources, NIAID, NIH: Genomic RNA from SARS-Related Coronavirus 2, Isolate USA-WA1/2020, NR-52285; SARS-Related Coronavirus 2, Isolate USA-WA1/2020, NR-52281; and Human Coronavirus, NL63, NR-470.

## AUTHOR CONTRIBUTIONS

S.S., D.A.F., and M.O. conceived and designed the study. S.S. and A.L. performed experiments and S.S. analyzed data. J.S., S.I.S., P.F., G.R.K., G.J.K. designed crRNAs. D.B. and C.S. assisted with BSL-3 work. J.S., S.I.S., P.F., G.R.K., G.J.K., D.C.J.S., T.Y.L. and J.A.D provided reagents. G.R.K., J.A.D., and M.O. obtained clinical samples. G.J.K., T.Y.L., and J.A.D. provided critical expertise and feedback. D.A.F. and M.O. supervised the study design and data collection. J.A.D., D.A.F., and M.O. secured funding. S.S. and D.A.F. wrote the manuscript, and all authors contributed feedback.

