## Supplementary Information for "Sensitive and multiplexed RNA detection with Cas13 droplets and kinetic barcoding"

SUPPLEMENTARY FIGURES

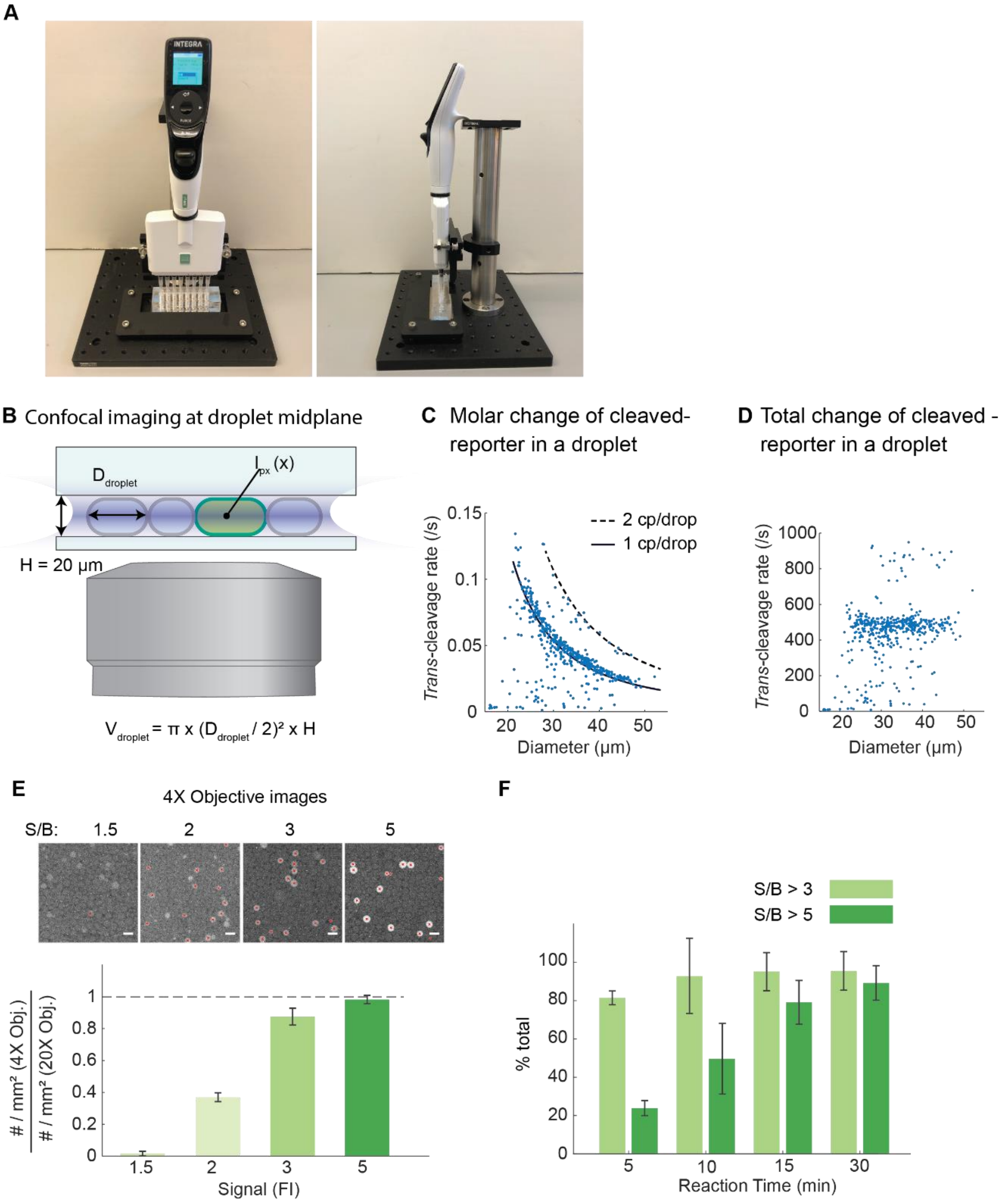


**Supplementary Fig. 1: Formation and imaging of Cas13 droplets**

a) Experimental setup for automated droplet formation consisting of an 8-channel automatic pipette, a custom pipette stand, and a tube rack.

b) Schematic depicting the geometry of flow cell and droplets, and confocal imaging of fluorescence signal (not drawn to scale). A 20X Water Immersion objective was used when droplet volume needed to be accurately determined. $D_{droplet}$ = droplet diameter, $V_{droplet}$ = droplet volume, $I_{px}(x)$ = fluorescence signal intensity of camera pixel in location x. Droplets larger than 20 µm deform to disks.

c) The slope of Cas13a signal accumulation in individual droplets containing crRNA 4 and 3.5 x $10^4$ copies/µL of SARS-CoV-2 RNA (N = 478 droplets). Fluorescence signal was obtained by integrating the pixel values within a droplet and converted to molar concentration based on a calibration using fully cleaved reporters. The solid and dotted black lines are obtained by performing a linear regression between the reaction velocity and the bulk-equivalent target concentration. It indicates the molar change of cleaved reporters for varying droplet size when a droplet contains one or two copies of RNA target.

d) As in c), but the *trans*-cleavage rate is shown in total copy number change of cleaved reporters in a droplet.

e) Comparison of detection sensitivity between 4X and 20X objective lenses. Top: Representative 4X images of positive droplets containing Alexa 488 dye, emulating a defined signal-to-background ratio (S/B). The red crosses mark positive droplets automatically detected from the custom image analysis routine. Scale bar = 50 µm. Bottom: The ratio of positive droplet counts quantified in a 4X versus 20X lens for the same droplets. Data are represented as mean ± SD of two replicates.

f) Fraction of positive droplets exhibiting S/B > 3 or 5 during reaction incubation in Fig. 1G and H, represented as mean ± SD of three replicates.

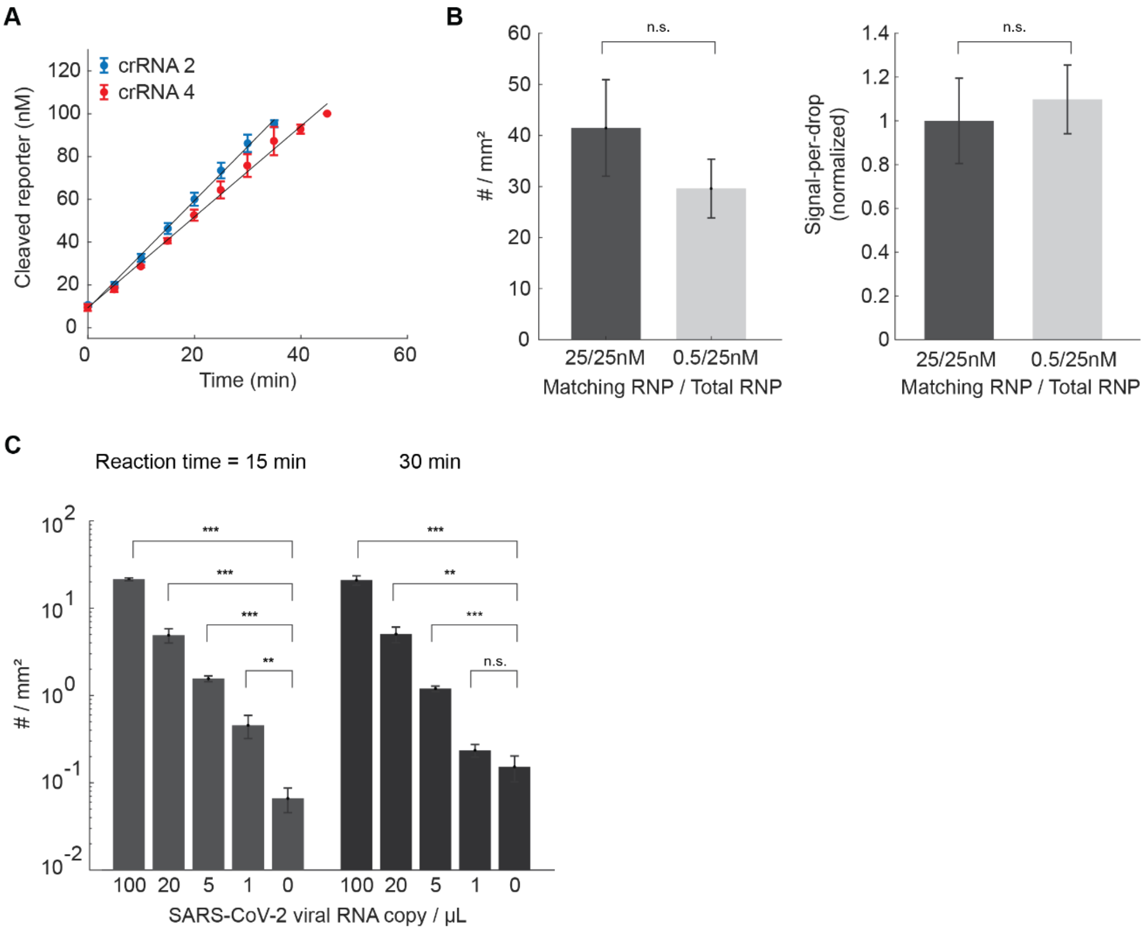


**Supplementary Fig. 2: crRNA combinations in the Cas13 droplet assay**

a) Bulk Cas13a reaction rate for 4 x $10^5$ copies/μL of *in-vitro* transcribed (IVT) N gene RNA was measured with crRNA 2 or 4 and represented as mean ± SD of three replicates.

b) Number of positive droplets per unit area (left) and fluorescence signal in individual droplets (right) measured from reactions containing 2.5 x $10^4$ copies/μL of IVT N gene RNA, together with either 25nM of an N gene-targeting crRNA (crRNA 4) only or combination of 0.5nM of crRNA 4 and 24.5nM of an E gene-targeting crRNA (crRNA 21). Data represents mean ± SD of three replicates. P-values determined based on a two-tailed *Student's* t-test are 0.14 (left), 0.63 (right). ns = not significant.

c) Number of positive droplets per unit area are quantified for the dilution series of externally quantified SARS-CoV-2 RNA after two different incubation times. Data are represented as mean ± SD of three replicates. P-values determined based on a two-tailed *Student's* t-test are 1.1e-6, 7.8e-4, 2.8e-5, 7.6e-3 for 15-minute reactions and 1.51e-4, 1.1e-3, 4.8e-5, 8.5e-2 for 30-minute reactions. ns = not significant, *p < 0.05, ** p < 0.005, ***p < 0.001.

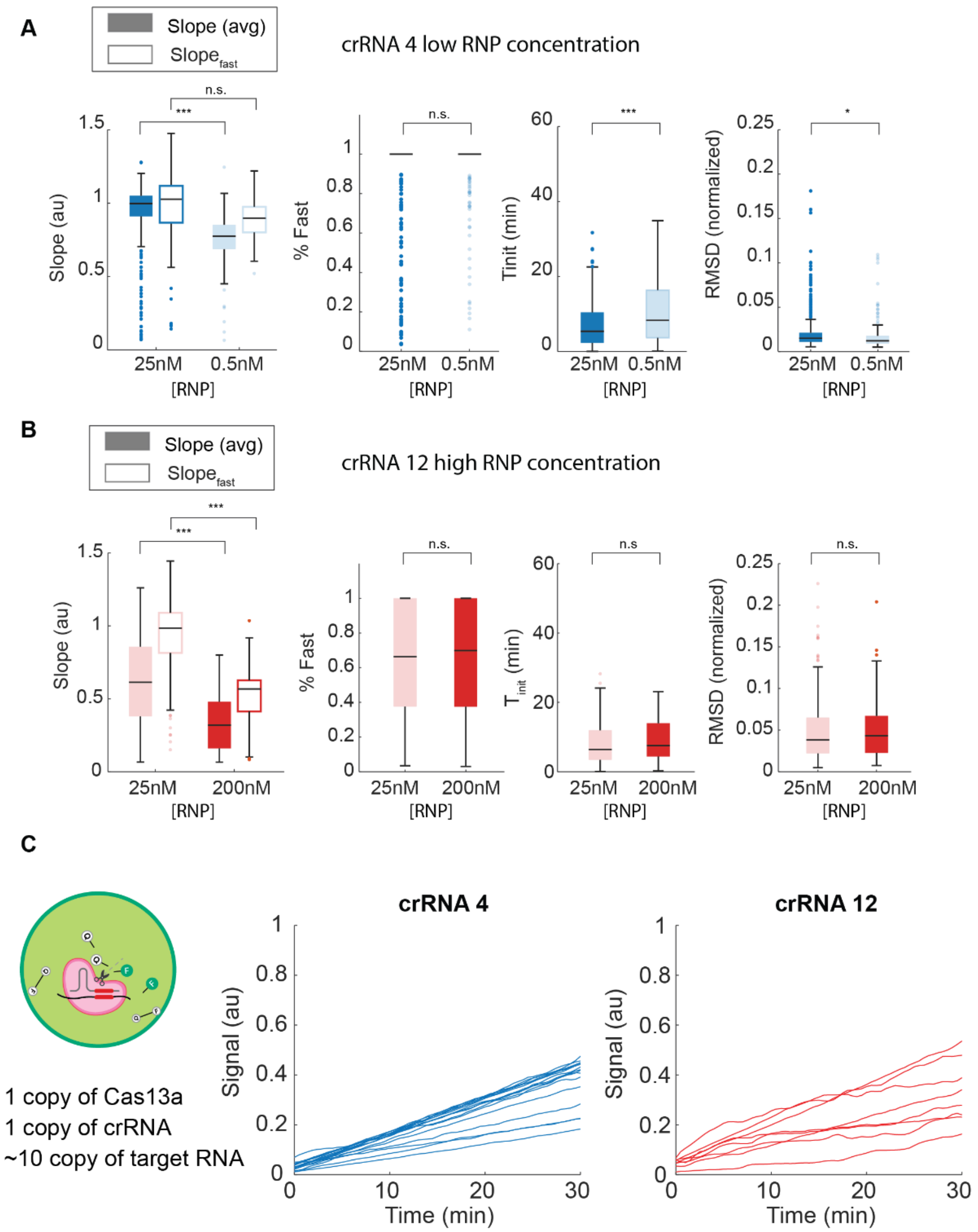

**Supplementary Fig. 3: Effect of RNP concentrations on the Cas13a reaction kinetic parameters.**

a) Effect of RNP concentration on Cas13a reaction kinetics using crRNA 4. Distribution of kinetic parameters are represented as box and whisker plots representing the median, the lower and upper quartiles, and the minimum and the maximum values, with outliers overlaid as individual data points. 395 and 190 individual 30-minutes-long trajectories are collected from two replicate experiments for each RNP concentrations. P-values determined from a two-tailed *Student's* t-test are 8.8e-18 and 0.13 (slope), 0.37 (% Fast), 9.2e-5 ($T_{init}$), and 1.1e-2 (RMSD). ns = not significant, *p < 0.05, ** p < 0.005, ***p < 0.001.

b) As in a, but for crRNA 12. We collected 311 and 102 individual trajectories from two replicate experiments for each RNP concentration. P-values determined from a two-tailed *Student's* t-test are 1.4e-17 and 6.1e-29 (slope), 0.69 (% Fast), 0.39 ($T_{init}$), and 0.81 (RMSD). ns = not significant, *p < 0.05, ** p < 0.005, ***p < 0.001.

c) Representative single Cas13a reaction trajectories measured in a droplet containing only single copies of Cas13a and crRNA (crRNA 4 or 12). Cas13a and crRNA were added in bulk concentrations of ~1 x $10^5$ copies/µL each (~1 copy/µL) and SARS-CoV-2 viral RNA was added in ~1 x $10^6$ copies/µL (~10 copies/uL), and a small fraction (<10%) of droplets showed positive reaction. Signal is total change of fluorescence in a droplet (arbitrary units).

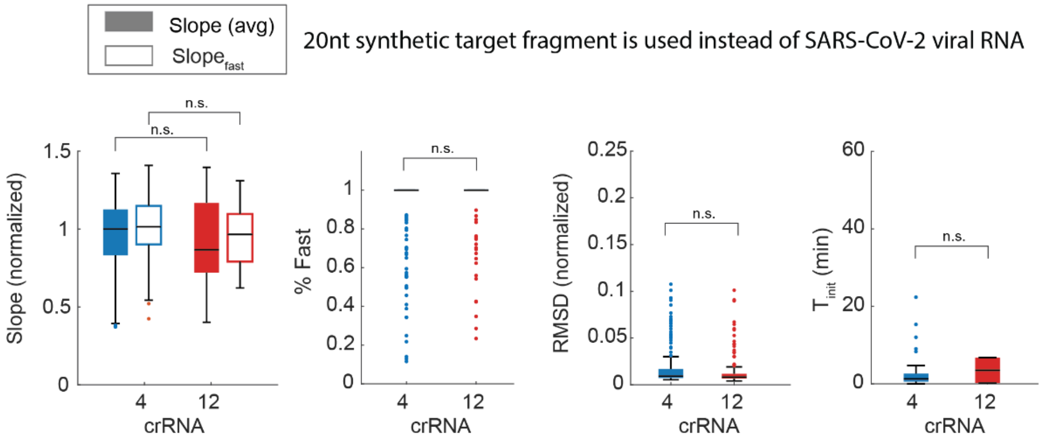


**Supplementary Fig. 4**: Effect of target RNA size on Cas13a reaction kinetics. Distribution of key parameters is represented as the box and whisker plots representing the median, the lower and upper quartiles, and the minimum and maximum values, with outliers overlaid as individual data points. We collected 230 and 129 individual 30-minutes-long trajectories from two replicate experiments for each crRNA. P-values determined from a two-tailed *Student's* t-test are 0.35 and 0.34 (slope), 0.61 (% Fast), 0.21 (RMSD), and 0.91 ($T_{init}$). ns = not significant.

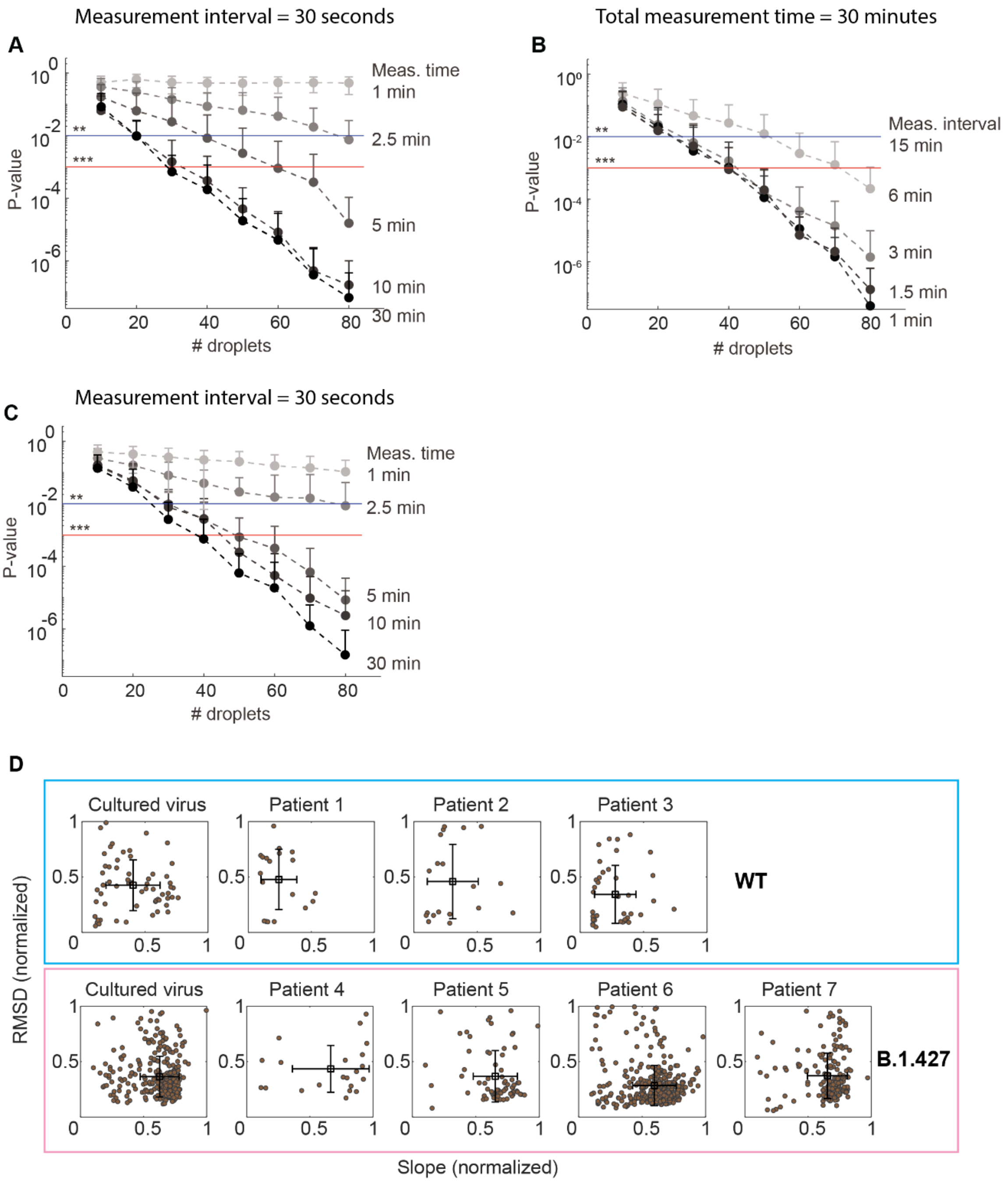


**Supplementary Fig. 5: Performance of the kinetic-barcoding approach with cultured viruses and clinical samples.**

a) Identification of HCoV-NL-63 or SARS-CoV-2 based on kinetic parameters of individual Cas13a reactions. Varying numbers of 30-minutes-long Cas13a trajectories are randomly selected from each condition in Fig. 4D and the difference between two groups are quantified as p-values based on a two-tailed *Student's* t-test. P-values are also determined for decreasing measurement times by taking the initial parts of Cas13a trajectories. Data shows mean ± SD of 50 repeated iterations. ** (the blue line) = $p < 0.005$, *** (the red line) $p < 0.001$.

b) P-values between HCoV-NL-63 and SARS-Co-2 for varying measurement intervals. Data shows mean ± SD of 50 repeated iterations.

c) Identification of the WT versus D614G IVT S gene based on kinetic parameters of individual Cas13a reactions. Two groups of reaction trajectories are selected from each condition in Fig. 4G and p-values were calculated for decreasing measurement times. Data shows mean ± SD of 50 repeated iterations.

d) Detection of SARS-CoV-2 variant from clinical samples based on kinetic-barcoding. Distribution of Cas13a kinetic parameters measured with RNA extracted from cultured wild-type (the blue box) or mutant viruses (the magenta box) as well as nasal/oral swab samples from people known to be infected with wildtype virus (the blue box) or the variant (the magenta box) based on sequencing. The black square and black cross indicate mean and standard deviation, respectively.

### Supplementary Table 1: Oligonucleotides used in this study

| Name | Sequence | Used in | Combination in Fig. 2D |
|---|---|---|---|
| crRNA 2 | GACCACCCCAAAAAUGAAGGGGACUAAAACGGUCCACCAAACGUAAUGCG | Fig. 2B-E, Supp Fig. 2C | 8, 17, 26 |
| crRNA 4 | GACCACCCCAAAAAUGAAGGGGACUAAAACUUUGCGGCCAAUGUUUGUAA | Fig. 1D-H, 2B-E, 3A-C, 3D, 3J-M, Supp Fig. 1C-F, 2A-C, 3A, 3C-D | 1, 8, 17, 26 |
| crRNA 11 | UAGACCACCCCAAAAAUGAAGGGGACUAAAACGCUUGUGUUACAUUGUAUGC | Fig. 3A-C, 3E, 3J-M | - |
| crRNA 12 | UAGACCACCCCAAAAAUGAAGGGGACUAAAACAAUUUGAUGGCACCUGUGUA | Fig. 3A-C, 3F, 3H-M, 4C-E, Supp Fig. 2C, 3B-D | 26 |
| crRNA D3 | UAGACCACCCCAAAAAUGAAGGGGACUAAAACAAACUACGUCAUCAAGCCAA | Fig. 2D-E, Supp Fig. 2C | 8, 17, 26 |
| crRNA D7 | UAGACCACCCCAAAAAUGAAGGGGACUAAAACCACAGUCAUAAUCUAUGUUA | Fig. 2D-E, Supp Fig. 2C | 8, 17, 26 |
| crRNA F1 | UAGACCACCCCAAAAAUGAAGGGGACUAAAACUCACACUUUUCUAAUAGCAU | Fig. 2D-E, Supp Fig. 2C | 8, 17, 26 |
| crRNA F6 | UAGACCACCCCAAAAAUGAAGGGGACUAAAACUGUAAGAUUAACACACUGAC | Fig. 2D-E, Supp Fig. 2C | 8, 17, 26 |
| crRNA H1 | UAGACCACCCCAAAAAUGAAGGGGACUAAAACUUAAUUGUGUACAAAAACUG | Fig. 2D-E, Supp Fig. 2C | 8, 17, 26 |
| crRNA H9 | UAGACCACCCCAAAAAUGAAGGGGACUAAAACCAGUUGUGAUGAUUCCUAAG | Fig. 2D-E, Supp Fig. 2C | 8, 17, 26 |
| crRNA A1 | UAGACCACCCCAAAAAUGAAGGGGACUAAAACUGAUAAGACCUCCUCCACGG | Fig. 2D-E, Supp Fig. 2C | 17, 26 |
| crRNA B1 | UAGACCACCCCAAAAAUGAAGGGGACUAAAACCAAAGCCUCAACACGUAGA | Fig. 2D-E, Supp Fig. 2C | 17, 26 |
| crRNA B2 | UAGACCACCCCAAAAAUGAAGGGGACUAAAACCAUGUUGAAACAAGUAACUC | Fig. 2D-E, Supp Fig. 2C | 17, 26 |
| crRNA C1 | UAGACCACCCCAAAAAUGAAGGGGACUAAAACUCAACGAUGUAAGAAGACUG | Fig. 2D-E, Supp Fig. 2C | 17, 26 |
| crRNA C2 | UAGACCACCCCAAAAAUGAAGGGGACUAAAACACACUAUCAACGAUGUAAGA | Fig. 2D-E, Supp Fig. 2C | 17, 26 |
| crRNA C8 | UAGACCACCCCAAAAAUGAAGGGGACUAAAACUAACCAUCCACUGAAUAUGU | Fig. 2D-E, Supp Fig. 2C | 17, 26 |
| crRNA H2 | UAGACCACCCCAAAAAUGAAGGGGACUAAAACAAACAGUAAGGCCGUUAAAC | Fig. 2D-E, Supp Fig. 2C | 17, 26 |
| crRNA H3 | UAGACCACCCCAAAAAUGAAGGGGACUAAAACUCCAAUUGUGAAGAUUCUCA | Fig. 2D-E, Supp Fig. 2C | 17, 26 |
| crRNA H5 | UAGACCACCCCAAAAAUGAAGGGGACUAAAACGACUAAAGCAUAAAGAUAGA | Fig. 2D-E, Supp Fig. 2C | 17, 26 |
| crRNA B3 | UAGACCACCCCAAAAAUGAAGGGGACUAAAACAAAAUUAUCGAAGCUUGCGU | Fig. 2D-E, Supp Fig. 2C | 26 |
| crRNA B8 | UAGACCACCCCAAAAAUGAAGGGGACUAAAACCAGUACAUCAAACGAAUUUG | Fig. 2D-E, Supp Fig. 2C | 26 |
| crRNA C12 | UAGACCACCCCAAAAAUGAAGGGGACUAAAACGGAAAUACAAAUGAUAUAAG | Fig. 2D-E, Supp Fig. 2C | 26 |
| crRNA E12 | UAGACCACCCCAAAAAUGAAGGGGACUAAAACAAUUUCCAUUUGACUCCUGG | Fig. 2D-E, Supp Fig. 2C | 26 |
| crRNA E4 | UAGACCACCCCAAAAAUGAAGGGGACUAAAACCAAUCUUUAAAGAGUCCUGU | Fig. 2D-E, Supp Fig. 2C | 26 |
| crRNA E7 | UAGACCACCCCAAAAAUGAAGGGGACUAAAACUUAAAUUGAUCUCCAGGCGG | Fig. 2D-E, Supp Fig. 2C | 26 |
| crRNA H6 | UAGACCACCCCAAAAAUGAAGGGGACUAAAACGAAGUAGACUAAAGCAUAAA | Fig. 2D-E, Supp Fig. 2C | 26 |
| crRNA H8 | UAGACCACCCCAAAAAUGAAGGGGACUAAAACCACCGGUGAUCCAAUUUAUU | Fig. 2D-E, Supp Fig. 2C | 26 |
| HCoV crRNA 6 | GACCACCCCAAAAAUGAAGGGGACUAAAACCCUCUCUGGUAGGAACACGC | Fig. 4C-E, Supp Fig. 4A-B | - |
| crRNA 6 | GACCACCCCAAAAAUGAAGGGGACUAAAACAAGAUCCUGAUAAAGAACAG | Fig. 4F-H, Supp Fig. 4C | - |
| crRNA 45 | GACCACCCCAAAAAUGAAGGGGACUAAAACCUUUCCAUACAACUUUUGUU | Fig. 4I, Supp Fig. 4D | - |
| crRNA 21 | GACCACCCCAAAAAUGAAGGGGACUAAAACAGCGCAGUAAGGAUGGCUAG | Supp Fig. 2B | |
| crRNA4 20nt target | GCACAGCAGAAAATCTCTGC**UUACAAACAUUGGCCGCAAA**CCACAG[1] | | |
| crRNA 12 20nt target | GCACAGCAGAAAATCTCTGC**UACACAGGUGCCAUCAAAUU**CCACAG[1] | | |
| PolyU Reporter | /56-FAM/rUrUrUrUrU/3IABkFQ/ | Fig. 3A, Supp Fig. 2A | |
| PolyU Reporter | /Alexa488/rUrUrUrUrU/3IABkFQ/ | Fig. 1-4, Supp Fig. 1-4 | |

[1] The oligonucleotide is consisted of DNA (underlined) followed by RNA. The 20-nt oligonucleotides complementary to the crRNA spacer sequence is indicated in bold.

**Supplementary Table 2: Clinical SARS-CoV-2 samples used in this study.**

| | GISAID ID | Ct Orf1ab | Ct S | S13I |
|---|---|---|---|---|
| **1** | CA-IGI-2633 | 14.85412 | 15.94596 | No |
| **2** | CA-IGI-2629 | 15.5905 | 16.90337 | No |
| **3** | CA-IGI-2626 | 14.43315 | 15.91746 | No |
| **4** | CA-IGI-2623 | 13.4006 | 14.69608 | Yes |
| **5** | CA-IGI-2631 | 14.64276 | 16.9693 | Yes |
| **6** | CA-IGI-2641 | 17.22917 | 19.7629 | Yes |
| **7** | CA-IGI-2628 | 16.36228 | 16.56808 | Yes |
